# Developing research knowledge through special study module: Notes from year 2 medical students

**DOI:** 10.1101/2021.01.12.21249687

**Authors:** S M Niazur Rahman, Nazmun Nahar Alam, Tanbira Alam

## Abstract

**Introduction:** Special Study Module is a mandatory research project implemented in some medical curriculum. For the successful application of evidence-based practice, physicians must understand research methodology before the appraisal of relevant evidence. The objective of this paper is to provide a brief overview of the effectiveness of student research modules in enhancing research understanding.

**Methods:** Second-year students of undergraduate medical programs were included in this study. At the beginning and after completion of literature review type research projects, the same student cohort completed a questionnaire comprising sections on perceived competency, attitude, and knowledge on research skills. Data from 163 participants who completed both pre and post-survey were analyzed.

**Results:** Paired t-test revealed significant differences in self-perceived knowledge, attitude, and assessed knowledge of the subjects before and after completing the research modules. Multivariate analysis also displayed a significant increase in self-perceived and assessed knowledge by carrying out the project. No significant association was exhibited by gender and ethnicity.

**Conclusions:** In summary, the research module improved students’ understanding of research methodology as well as the structure of scientific communication. Further studies to assess the effect of various types of research modules in different study populations are warranted.

## Introduction

Research has been the driving force in initiating new directions on the improvement of medical science. The involvement of medical students in research activities is imperative to develop critical thinking, efficient literature-searching, and publishing to contribute to medicine.^1^ Despite the evidence-based practice being core standards for all health professionals, relatively few of them are involved in the research. ^2^

Special Study Modules (SSM) is a research study program implemented in medical schools, aimed to develop students’ research skills. ^3, 4^ Of many, one vital purpose of SSM is the intellectual development of students through exploring in-depth subjects of their choice. The objective of this study was to describe the effectiveness of the SSM project in learning the basic research methodology and scientific writing structure by the second-year medical students. Additionally, this project referred to the self-perceived competency and attitudes of students towards research.

## Methods

### SSM research project

The SSM research in the preclinical phase is conducted from the beginning of year 2 until 6 weeks before the second professional exam. All 2nd-year students are divided into small groups and assigned with a lecturer as a supervisor who had research experience to guide them along the way. Usually, there are 4-6 students in a group, and they are allowed to choose the area of research they are interested in, from different aspects of medicine ranging from basic to the clinical topic. Thus, students are assigned to the lecturers involving all the departments of the institute. Students are encouraged to engage in an in-depth study of something related to medicine and health care that interests them but is not covered in detail in the core curriculum. SSM in the preclinical year is designed with the primary focus on literature review, through which learners can understand the structure of a journal article in detail, and improve their concept of overall research methodology. In clinical years, they undertake SSM of practical research type; designing a study followed by data acquisition and analysis and final report writing. Our project on SSM in the second year is also termed as library SSM.

### Study design

All students enrolled in the year 2 program (*n* = 200) were invited to participate in this study and voluntary informed consent was obtained from each of them. Ethics approval was granted by the relevant university human research ethics committee (approval number: AUHEAC/FOM/2016/08). A pre-post evaluation was used to measure changes in knowledge about research methodology and scientific writing. Self-perceived research knowledge and attitudes towards research were also noted through the pre-post type survey. Prior to the commencement of the SSM project, all participants completed a research skills questionnaire comprising of 20 items designed to test their knowledge on research methodology and scientific article. Students were then asked to complete the same survey on completion of the project. The survey required participants to rate their self-perceived competency on item-specific research on a Likert scale of 1 to 5, with 1 representing poor competence and 5 the highest. In addition, attitudes by asking questions on their general feelings towards research such as motivation, interest, inspiration, excitement and usefulness were assessed on a scale of 1 to 7. Only students who completed both pre and post surveys were included in the analysis to assess the changes in knowledge, perceived competency and attitude. Descriptive statistics were used to describe demographics. A paired t-test was done to assess the differences in knowledge of the participants before and after the module. Multivariate analysis was used to assess if there was a difference between self-perceived competence in research skills and assessed knowledge of research, prior to and following completion of the SSM. Significance was considered at p < 0.05.

## Results

There were 191 respondents recruited at the beginning of the study. However, 163 students responded to the post-survey, hence, they were included for further analysis. The average age of 163 participants was 20 years, which included 68 (41%) females, and 80 (49%) belonged to Chinese ethnicity. The demographic information is displayed in Table 1.

**Table 1.** Demographic data of the respondents

| Features | Respondents: n = 163 |
| --- | --- |
| Age | 20.44±0.84 |
| Gender | 68 Male, 95 Female |
| Ethnicity | 80 Chinese, 77 Indian, 06 Others |

A paired-samples t-test was conducted to assess the effect of the SSM project on the perceived competency, attitude and knowledge of the respondents towards research. There were statistically significant differences in self-perceived competency and attitude as shown in Table 2.

**Table 2.** Paired sample statistics of self-perceived competency and attitude

|  |  | Difference of Mean | SD | t | Sig (2-tailed)<br>*p* |
| --- | --- | --- | --- | --- | --- |
| Self-perceived competency | Pre SSM | 0.74 | 0.87 | 10.87 | 0.00 |
|  | Post SSM |  |  |  |  |
| Attitude | Pre SSM | 0.45 | 1.13 | 5.04 | 0.00 |
|  | Post SSM |  |  |  |  |

df (degree of freedom): 162. SSM, Special Study Module.

There was a statistically significant difference in assessed knowledge score between pre SSM (Mean 11.63 ± SD 2.33) and post SSM (Mean 12.02 ± SD 2.29) phase, t (162) = −2.151, p<0.03 (2-tailed). The mean increase in knowledge score was −0.387 with a 95% confidence interval ranging from −0.74 to −0.03.

Multivariate analysis by a one-way repeated measure of ANOVA revealed, there was a significant effect (Wilk’s Lambda= 0.97, F (1,162) = 4.26, p<0.03) of SSM project on the perceived knowledge and actual scored knowledge by the respondents. These results statistically suggest a moderate effect size (multivariate partial eta squared = 0.03).

There was no significant relationship displayed by the gender and ethnic groups of the respondents analyzed by the chi-square test.

## Discussion

Our study focused on the effectiveness of the SSM project in enhancing research skills. The main purpose of SSM is having hands-on experience in the research methodology, reading and reviewing literature helps learners to do so effectively. Moreover, this process helps students to learn how to present their understanding of primary biomedical literature in their own words and figuring out the structure of a scientific article.

In recent days, many medical schools incorporate mandatory courses on research which could allow students to carry out all the steps of a research project from conception to final report writing, thereby narrowing the gap between theory and practice. ^5^ In addition, SSM projects encourage students to pursue their future research activities and enter an academic and research career. ^6^ Since the topics are drawn from all areas of medical science, SSM also assists in the development of professional, transferable skills for life-long learning in medicine. ^7^ General Medical Council (GMC) stipulated that SSMs “must be an integral part of the curriculum, enabling students to demonstrate mandatory competencies, while allowing choice in studying an area of particular interest to them”. ^8^

Research studies are part of the medical curriculum in developed countries such as the USA, UK, and Canada. Expert supervision, financial support and summer internship programs provided for the students in such countries encourage them to develop, design and carry out their research projects on time and broaden their skills. ^9^ Researchers suggest that students are more likely to be benefited when they are involved in research; they achieved more sophisticated levels of intellectual development. ^10^ Many international journals including BMJ, Lancet, PLoS medicines have been including student sections regularly, thus motivating students’ perspectives about the critical issues regarding health care ^1^. Though research opportunities are limited in most South Asian countries, the importance of the student research activities is recognized very well, and it is incorporated in the curriculum. ^11^ There is an increasing emphasis among allied health professionals to link research evidence and clinical practice thereby enhancing the clinical decision-making process. ^12^

The negative attitudes of medical students toward research are an obstacle to learning associated with poor performance in research. ^13, 14^ Our results are suggestive of significant shifting of the students’ attitude towards positivity (p < 0.00) after completing the SSM project.

Most of the respondents from Canadian medical schools ^14^ have pointed out the insufficient training on research methodology and critical appraisal of scientific literature, as a barrier in research. The results of this study have demonstrated significant improvement in understanding (p < 0.00) on research methodology as well as scientific articles.

In a previous study, self-perceived research skill was used to reflect on learning rather than actual assessment. ^15^ However, only self-assessment of skills has been acknowledged as a major limitation. ^16^ As measuring actual knowledge in research is crucial, we have followed our cohort longitudinally and used validated measures to assess the effectiveness of research involvement in the undergraduate learning experience.

Our study was initiated by a pre-survey followed by an intervention through a designed SSM research project and finally concluded with the post-survey, all by the same respondents. Hence, we could consider the data as strong evidence of assessing research effectiveness.

The outcome of this study is limited concerning the study sample; they are from one university and there was a drop out of 12% of subjects from the pre to post-survey. Moreover, those who completed both surveys perceived as slightly more confident, engaged and perhaps had the best learning outcomes.

To the best of our knowledge, this study is the first to evaluate the effectiveness of a preclinical SSM in enhancing knowledge, perceptions and attitudes toward research among medical students in the Southeast and South Asian region. We not only addressed an essential yet neglected issue in our region but also attempted to comprehensively assess the importance of the research module to be included in the curriculum. We believe, our efforts would lead medical students more towards the research. Future research could explore the comparison in multiple groups through different approaches of SSM modules assigned to each cohort.

## Conclusions

In the era of continuous professional development and evidence-based medicine, physicians need at least a basic understanding of research methodology, so as to the interpretation of published research findings. This approach will enable them to carry out their research projects. It is a vital responsibility of doctors to train themselves to an extent, at which they are capable of discerning good from bad research studies. By doing so, medical practitioners will be able to verify whether the conclusions of a study are valid and what are the constraints of such studies. From the above context, we can conclude that performing research module training is obligatory and our present study strongly supports the incorporation of such modules in the early years of the medical curriculum.

## Disclosure

No conflicts of interest, financial or otherwise, are declared by the authors.

## Data Availability

Yes

## Acknowledgement

We acknowledge the contribution of Dr. Nazma Farhat during the data collection of this study.

## Author Contributions

S.M.N.R., N.N.A., and T.A. conceived and designed research; T.A. analyzed data; S.M.N.R., N.N.A., and T.A. interpreted results of experiments; S.M.N.R. prepared tables; S.M.N.R. and N.N.A. drafted the manuscript; S.M.N.R., N.N.A., and T.A. edited and revised manuscript; S.M.N.R., N.N.A., and T.A. approved the final version of the manuscript.

## Author Notes

S.M.N.R. and T.A. were in full time academic position at the faculty of medicine, AIMST University, while the study was done.

## References

1. Basnet B, Bhandari A. Investing in medical student’s research: Promoting future of evidence based medicine in Nepal. Health Renaissance. 2013;11(3):297–300.

2. Stephens D, Taylor N, Leggat SG. Research experience and research interests of allied health professionals. Journal of Allied Health. 2009;38(4):107E–11E.

3. Salam A, Hamzah JC, Chin TG, Siraj HH, Idrus R, Mohamad N, et al. Undergraduate medical education research in Malaysia: time for a change. Pakistan journal of medical sciences. 2015;31(3):499.

4. Guner GA, Cavdar Z, Yener N, Kume T, Egrilmez MY, Resmi H. Special-study modules in a problem-based learning medical curriculum: An innovative laboratory research practice suppporting introduction to research methodology in the undergraduate curriculum. Biochemistry and Molecular Biology Education. 2011;39(1):47–55.

5. Aslam F, Shakir M, Qayyum MA. Why medical students are crucial to the future of research in South Asia. PLoS Med. 2005;2(11):e322.

6. Solomon SS, Tom SC, Pichert J, Wasserman D, Powers AC. Impact of medical student research in the development of physician-scientists. Journal of Investigative Medicine. 2003;51(3):149–56.

7. Hassan SL. Applying research-based learning in medical education through the route of special study modules: Notes from the UK. SA-eDUC JOURNAL. 2013;10(1):1-26,1810-6293.

8. Council GM. Tomorrow’s doctors: outcomes and standards for undergraduate medical education. Manchester, UK: General Medical Council. 2009.

9. Ogunyemi D, Bazargan M, Norris K, Jones-Quaidoo S, Wolf K, Edelstein R, et al. The development of a mandatory medical thesis in an urban medical school. Teaching and learning in medicine. 2005;17(4):363–9.

10. Healey MJ, Roberts J. Engaging students in active learning: Case studies in geography, environment and related disciplines: Geography discipline network, University of Gloucestershire; 2004.

11. Shankar P, Chandrasekhar T, Mishra P, Subish P. Initiating and strengthening medical student research: Time to take up the gauntlet. Kathmandu University medical journal (KUMJ). 2006;4(1):135.

12. Grimmer-Somers K. Incorporating research evidence into clinical practice decisions. Physiotherapy Research International. 2007;12(2):55–8.

13. AlGhamdi KM, Moussa NA, AlEssa DS, AlOthimeen N, Al-Saud AS. Perceptions, attitudes and practices toward research among senior medical students. Saudi Pharmaceutical Journal. 2014;22(2):113–7.

14. Siemens DR, Punnen S, Wong J, Kanji N. A survey on the attitudes towards research in medical school. BMC medical education. 2010;10(1):4.

15. Davidson ZE, Palermo C. Developing research competence in undergraduate students through hands on learning. Journal of Biomedical Education. 2015;2015.

16. Van Der Vleuten CP. The assessment of professional competence: developments, research and practical implications. Advances in Health Sciences Education. 1996;1(1):41–67.

